# Emerging Multi-Source Transmission of SFTS Virus on a Remote Japanese Island: A One-Health Perspective

**DOI:** 10.1101/2025.07.02.25330427

**Authors:** Ryosaku Oshiro, Motoki Ihara, Catarina Harumi Oda Ibrahim, Kosuke Matsui, Xayavong Dalouny, Nhung Hong Pham Vu, Tomomi Kurashige, Qiang Xu, Shangfan Hu, Mya Myat Ngwe Tun, Yumika Takaki, Hikaru Ogushi, Daichi Setoguchi, Shinichi Katsuoka, Tatsuki Murakami, Chikara Matsumoto, Mitsuru Hattori, Naoko Hattori, Sho Miyamoto, Yusuke Sakai, Tadaki Suzuki, Hirotomo Yamanashi, Hiroyuki Murota, Takahiro Maeda, Kouichi Morita, Akira Yoshikawa, Yuki Takamatsu

**Author notes:** **Correspondence author:** Yuki Takamatsu, Department of Virology, Institute of Tropical Medicine, Nagasaki University, 1-12-4 Sakamoto, Nagasaki 852-8523, Japan. These authors contributed equally to this work (Ryosaku Oshiro, Motoki Ihara).

## Abstract

Tick-borne viral infections, including Severe Fever with Thrombocytopenia Syndrome (SFTS), pose a growing global public health threat owing to their potential for rapid spread and high mortality rates. We report a localized outbreak of SFTS virus (SFTSV) affecting both humans and cats on a remote island in Nagasaki, Japan, between April and May 20xx, comprising four feline and four human cases. Notably, no direct contact was observed between cats and humans. Four animals and one patient succumbed to the illness. Genomic analyses, conducted in collaboration with local health authorities, revealed a high similarity to previously reported strains in Nagasaki. Haplotype network analysis suggested multiple sources and complex transmission pathways with evidence of diverse viral introduction events. The inconsistency across viral segments underscores the complexity of virus circulation. This outbreak underscores the critical importance of a One Health approach—integrating human, animal, and vector surveillance—to effectively monitor, understand, and mitigate the public health impact of tick-borne viruses, which are an increasingly significant threat in endemic and emerging regions worldwide.

## Introduction

Tick-borne viruses are increasingly recognized as a significant and growing threat to global public health. These pathogens, transmitted primarily by ticks, can cause severe and often emerging infectious diseases, with some viruses—such as Crimean-Congo hemorrhagic fever virus, tick-borne encephalitis virus, and Severe Fever with Thrombocytopenia Syndrome virus (SFTSV)—resulting in high mortality rate^1-4^. The geographic distribution and incidence of tick-borne viral infections are expanding due to factors such as climate change, increased human mobility, and changes in land use, which facilitate tick proliferation and human exposure. Furthermore, the involvement of multiple hosts—including humans, livestock, and wildlife—as well as the complex ecological interactions make effective prevention and control of these viruses particularly challenging, highlighting the urgent need for enhanced surveillance, research, and international cooperation to mitigate their impact on public health worldwide.

Severe Fever with Thrombocytopenia Syndrome (SFTS) is an emerging tick-borne viral disease of significant public health concern, attributed to *Dabie bandavirus*, a member of the *Phenuiviridae* family ^5^. This virus has a tripartite segmented RNA genome (L, M, and S), each of which is critical for infectivity and pathogenicity. The L segment encodes RNA-dependent RNA polymerase, essential for viral replication, while the M segment encodes glycoproteins vital for host cell entry and viral assembly. The S segment plays a dual role, encoding a nonstructural protein involved in immune evasion and the nucleocapsid protein (NP), which protects viral RNA by forming ribonucleoprotein complexes, thereby protecting it from degradation by host immune responses and nucleases ^6 7^. Understanding the molecular biology of SFTSV is crucial for developing effective diagnostics, vaccines, and therapeutic strategies to combat this high-mortality disease.

The first human case was reported in China in 2009 ^8^, and subsequent cases have been documented in Japan since 2013 ^9^, with approximately 50 to 100 cases annually, occasionally rising to over 100. While primarily transmitted by the tick *Haemaphysalis longicornis*, human-to-human transmission ^10^ and direct transmission from animals, such as cats, have also been reported ^11^, highlighting the complex epidemiology of SFTS. Clusters of infections within households ^10 12 13^ or veterinary clinics suggest clear links between human and animal hosts ^11^. However, to date, there are no reports of large, unlinked outbreaks involving multiple animals or humans confined to a specific area over a short period, raising concerns regarding the potential for undetected or emerging outbreaks. Understanding these transmission dynamics is critical for implementing effective public health interventions and preventing wider spread.

In Japan, laboratory diagnostics of suspected SFTS cases in cats and dogs, based on detection of viral genes and antibodies, have revealed that Nagasaki has the highest number of feline cases nationwide ^14^. However, epidemiological data from Nagasaki ^15^ do not show a clear geographic clustering of infected cats, and no cases among household members or nearby residents have been confirmed. From April to May 2024, four cats and four humans were reported to be infected within the same area of the Goto Islands, Nagasaki, raising concerns about local transmission dynamics. In response, we conducted a collaborative investigation with local hospitals and the Nagasaki Prefectural Institute of Environment and Public Health to better understand this outbreak. These findings highlight the complexity of SFTS transmission and underscore the need for integrated surveillance of human and animal populations.

### Concurrent SFTS infection

Clinical information was collected through collaboration with Nagasaki Goto Chuo Hospital and Matsumoto Veterinary Clinic. The clinical courses of the eight cases are summarized in Figure 1. The first case involved a cat (cat 1) that was referred to a veterinary clinic on day 0 (April, 20xx), exhibiting fever, anorexia, and jaundice. SFTSV RNA was detected in its blood on day 3. A second cat (cat 2), examined on day 10 with similar symptoms, also tested positive for SFTSV RNA in blood. On the same day, a third cat (cat 3) succumbed to the virus, which was subsequently identified in its oral swab. The fourth cat (cat 4) presented with comparable symptoms and was diagnosed with SFTS in mid-May. Regarding human cases, two individuals (humans 5 and 6) began developing symptoms, such as fever and malaise, in early May and sought medical care, where SFTS was confirmed. A third patient (human 7) experienced symptom onset shortly after, but unfortunately died in mid-May; post-mortem testing detected SFTSV in the blood. The eighth case (human 8), presenting with fever and headache, was diagnosed in mid-May as well.

**Figure 1.**
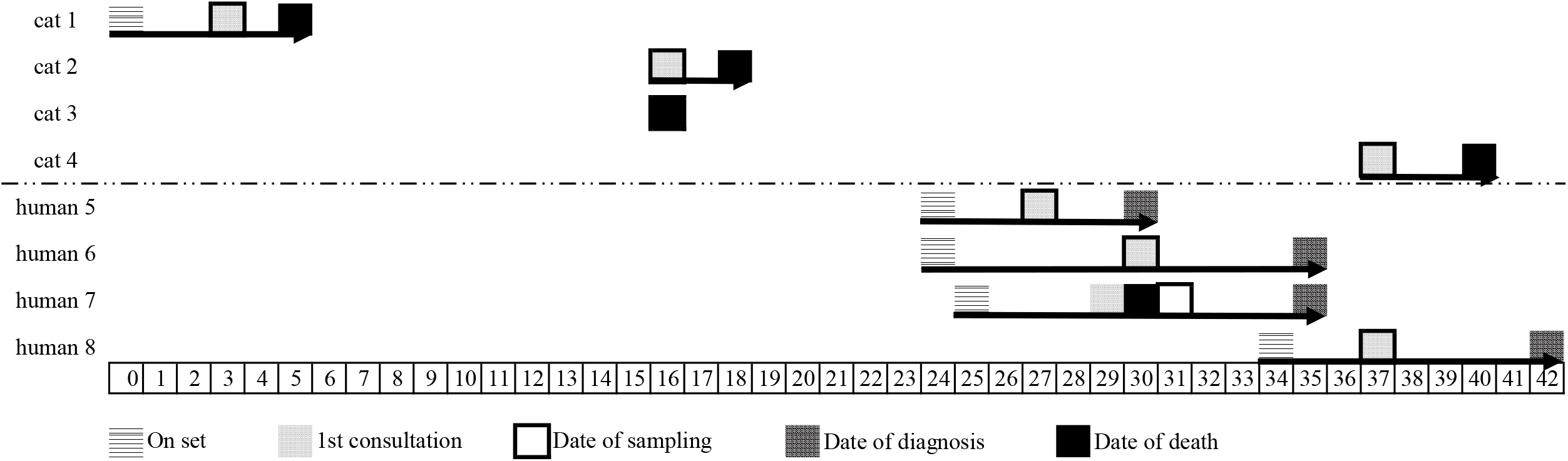
Timeline of eight cases (four cats and four humans). The onset days for cats 2, 3, and 4 are unknown. Cat 3 was found dead and diagnosed with SFTS from an oral swab sample.

No direct contact was observed between the four cats and the four humans. However, it was reported that the cats were domestic and freely roamed outdoors. Importantly, the patients were not owners of these cats. Three of them (humans 5, 6, and 8) engaged in outdoor activities, such as mowing grass and working in fields. Although human 7 died, he had no history of tick contact and kept healthy cats as pets.

Clinical presentations and laboratory results are summarized in Table 1. Human cases exhibited typical SFTS symptoms, including fever, anorexia, malaise, neurological symptoms, and hemoptysis. The cats showed similar signs of fever, lethargy, jaundice, and hemoptysis, which is consistent with previous reports ^16^. The high lethality observed in this outbreak is particularly alarming; all infected cats succumbed to the disease, highlighting its extreme virulence. In humans, the case fatality rate has been reported to be approximately 27% ^17^ whereas in cats, the mortality is approximately 62% ^18^.

**Table 1.**
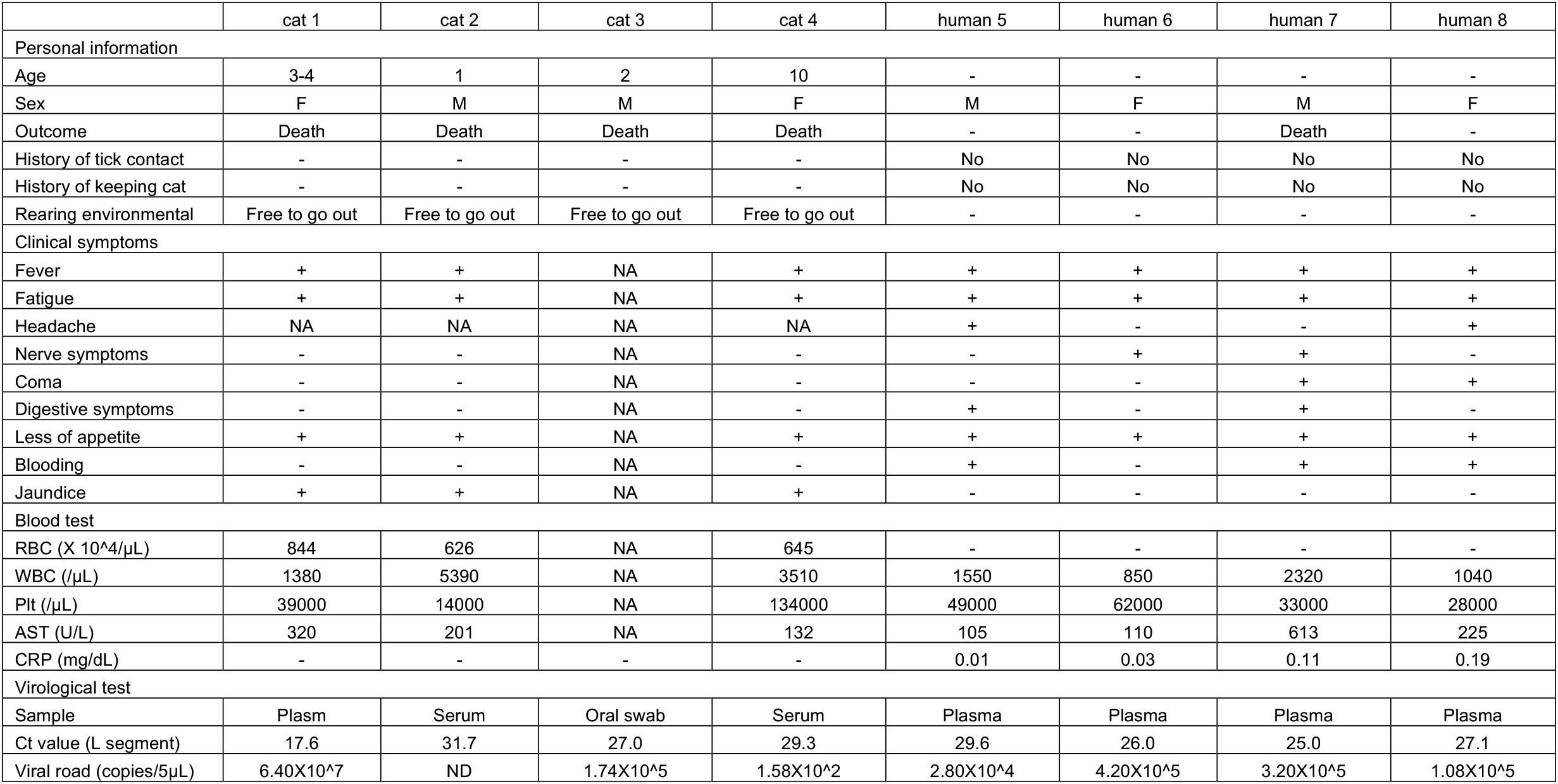
Clinical symptoms and hematologic and diagnostic results from eight cases. NA, not applicable; ND, not detected.

### Phylogenetic analysis

At the Nagasaki Prefectural Institute of Environment and Public Health, Ct values were determined via reverse transcription-quantitative PCR (RT-qPCR), targeting the L segment of SFTSV. Viral loads were also assessed at Nagasaki University using RT-qPCR targeting the NP gene in the S segment (Table 1). Detailed experimental protocols are provided in the supplementary appendix. No correlation was observed between viral load and clinical outcomes. Notably, the virus was undetectable in the S segment of cat 2, likely due to RNA degradation caused by temporal delays and repeated freeze-thaw cycles. Given the higher sensitivity of next-generation sequencing (NGS), the complete viral genome was successfully identified using NGS.

Whole-genome sequencing was performed via amplicon sequencing and deposited in the DNA Data Bank of Japan (Accession numbers: LC864283–LC864306). Phylogenetic analysis, incorporating eight samples alongside previously published sequences (Figure 2), revealed that all eight strains belonged to genotype B2, consistent with strains previously reported in Nagasaki ^15^. This indicates a localized viral population. Amino acid analysis across the three segments (Table 2) identified minor variations. In the L segment, most samples contained glutamine and lysine at positions 58 and 1705, respectively, while human 8 exhibited leucine and arginine at these sites. In the same segment, cat 4 carried lysine at position 1189, contrasting with arginine in other samples. In the M segment, while most samples were conserved, human 7 uniquely harbored isoleucine, serine, proline, histidine, and threonine residues at positions 232, 341, 462, 467, and 468, respectively. Human 7 was the only fatal case to date; however, no direct link between these amino acid substitutions and virulence could be established. In the S segment, cat 1 displayed valine at position 150, whereas the other samples possessed glycine at this site. Overall, these findings suggest that amino acid mutations are unlikely to fully explain the severity or outbreak dynamics of this SFTS cluster.

**Table 2.** Comparison of amino acids in the eight samples. The colored positions indicate different amino acids from the other samples.

|  |  | cat 1 | cat 2 | cat 3 | cat 4 | human 5 | human 6 | human 7 | human 8 |
|---|---|---|---|---|---|---|---|---|---|
| L | 58 | Q | Q | Q | Q | Q | Q | Q | L |
|  | 1189 | R | R | R | K | R | R | R | R |
|  | 1705 | K | K | K | K | K | K | K | R |
| M | 232 | V | V | V | V | V | V | I | V |
|  | 341 | P | P | P | P | P | P | S | P |
|  | 462 | L | L | L | L | L | L | P | L |
|  | 467 | Y | Y | Y | Y | Y | Y | H | Y |
|  | 468 | M | M | M | M | M | M | T | M |
| S | 150 | V | G | G | G | G | G | G | G |

**Figure 2.**
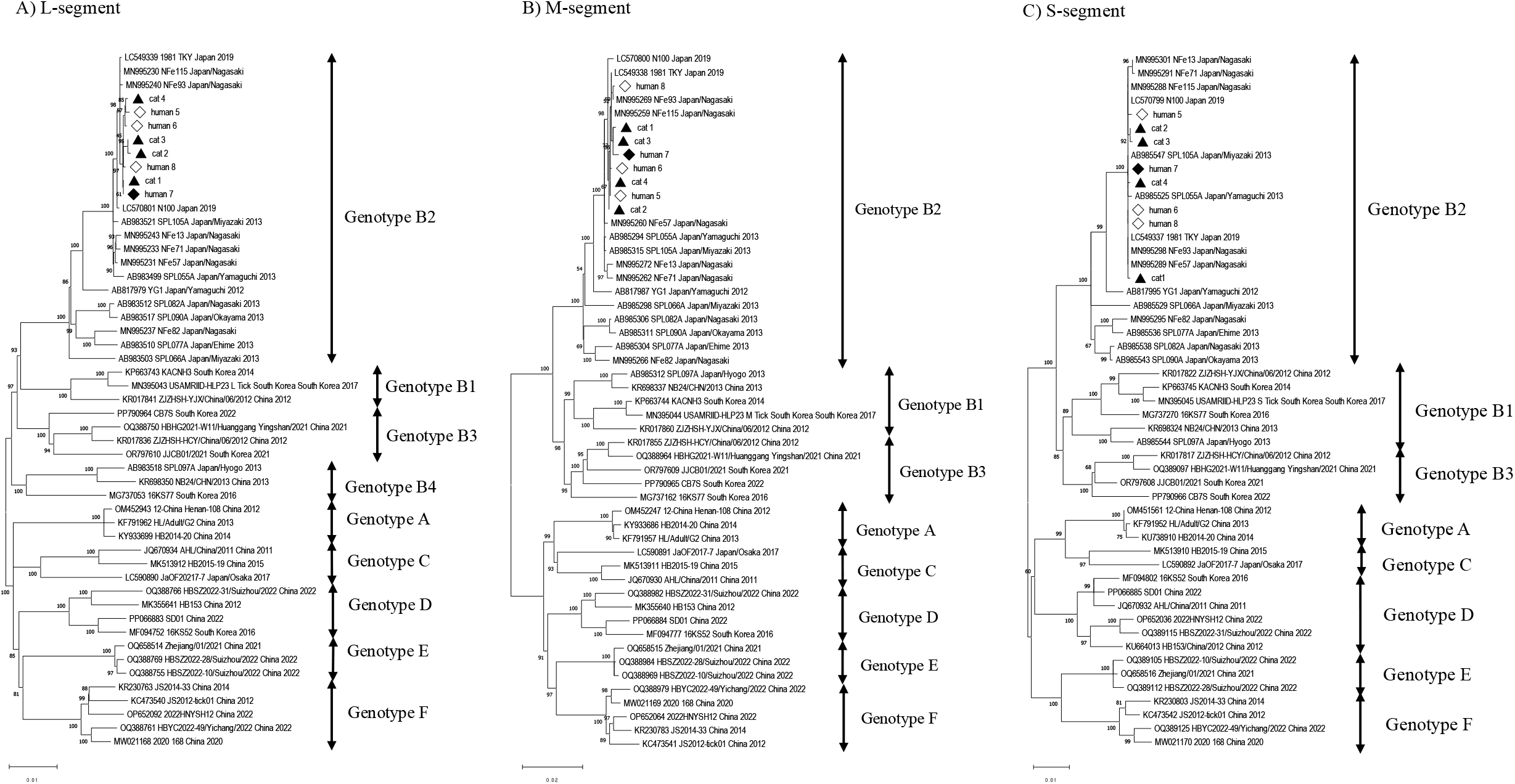
Phylogenetic analysis of SFTSV strains obtained from eight cases and previously reported sequences. Maximum likely food method, bootstrap 1000 times. A) L, B) M, and C) S segments. (▲) means dead cat, (♢) means saved human, (◆) means dead human.

### Haplotype network analysis

Haplotype network analysis is a valuable tool for elucidating molecular epidemiology and phylogeographic relationships within infectious disease outbreaks. Although its use has been well documented in studies on viruses such as SARS-CoV-2 ^19 20^, to date, this methodology has not been applied to SFTS. Therefore, we conducted haplotype network analysis on eight viral genomes obtained from our samples, alongside six previously reported strains from Nagasaki, to investigate potential transmission pathways (Figure 3). In the L segment, human 7 and cat 1 shared the most closely related haplotypes, differing only by a single nucleotide. Cats 2 and 3 differed by four bases, while cat 4 and human 6 differed by six bases. Similarly, human 7 and strain NFe115 also differed by six bases. In the M segment, cat 4 and human 5 shared the same haplotype (Haplotype I), which was also closely connected to cat 2, differing by only one base. Haplotype I further linked to human 6 and strain NFe115, with a two-base difference. Notably, in the L segment, the closest sequences—cat 1 and human 7—differed by nine bases, with cat 3 positioned between them in the network. Overall, the analysis suggests potential transmission links from cat 3 to cat 1, human 7, and human 8, and from Haplotype I to cat 2 and human 6 in the M segment. In the S segment, cats 2 and 3 clustered together with Haplotype II, whereas humans 6, 7, 8, and strains NFe57, NFe93, and NFe115 shared Haplotype III. Haplotype III was closely related to cats 1 and 4, as well as Haplotype II, differing by only one base in the S segment. Human 5 and Haplotype III were linked with a two-base difference. Across the segments, some cases showed direct phylogenetic links—such as between cat 1 and human 7 in the L and S segments—while others involved indirect connections via intermediate hosts or strains. However, no consistent pattern of haplotype correlation was observed across all segments. These findings suggest complex transmission dynamics that may involve vectors such as ticks or other animals facilitating viral spread. The observed heterogeneity across segments underscores the possibility of multiple introduction events and complex circulation patterns within the outbreak area.

**Figure 3.**
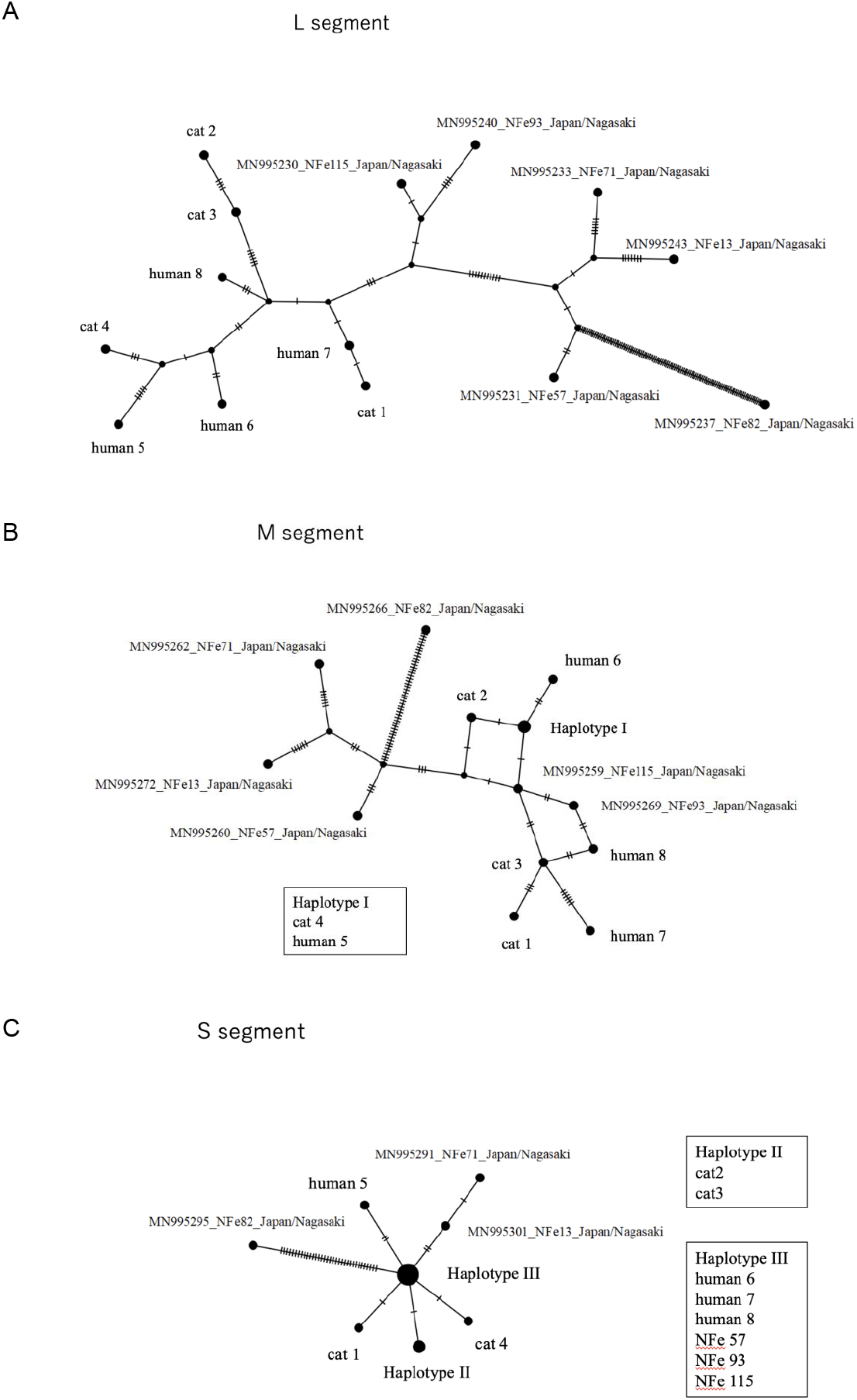
Haplotype network analysis using the minimum spanning network method for eight cases and six previously reported sequences from Nagasaki. A) L, B) M, and C) S segments. Hatch marks between the strains indicate the number of mutations. The strains in the square frame indicate the same haplotype. (▲) means dead cat, (♢) means saved human, (◆) means dead human.

### Challenges and One Health Strategy

This investigation faced key limitations. Notably, ecological samples and environmental conditions including ticks could not be collected during the epidemic period. The observed viral heterogeneity indicates the likelihood of multiple SFTSV sources contributing to the outbreak. Given the peak tick activity season, vector ticks likely played a role; however, delayed sampling hampered timely collection. Fortunately, no further cases have been reported in this area, indicating a potential transient event. Privacy constraints further limited the data collection, which could have enhanced the analysis.

Building on these findings, we established a one-health collaborative team—including regional authorities, virologists, tick ecologists, clinicians, and modelers—to proactively prevent future outbreaks. Evidence suggests that transiently aggregated infected ticks contribute to multiple human and animal cases; however, the precise origin of SFTSV remains unclear. Despite ongoing tick surveillance, no virus was detected—likely due to temporal limitations—despite previous extensive tick collection within Nagasaki Prefecture that also yielded no SFTSV ^21^.

Recent outbreaks in China, where SFTSV was identified in dozens of foxes across multiple farms, underscored the complex multi-source transmission dynamics ^22^. These findings highlight the critical need for timely, coordinated surveillance and a comprehensive One Health approach to decipher viral ecology. Future efforts will focus on identifying key reservoir hosts through serological analysis of wild and domestic animals, aiming to uncover infection sources and reduce outbreak risks.

## Conclusion

This study confirms the occurrence of a significant SFTSV epidemic affecting both humans and cats on a remote Japanese island. Despite the absence of direct contact between cats and humans, genomic analysis demonstrated closely related viral strains circulating concurrently within both populations over a two-month period. Incongruence across viral segments suggests multiple introductions and complex transmission pathways, underscoring the critical importance of the One Health approach. Integrating human, animal, and vector surveillance is essential for accurate risk assessment and development of effective intervention strategies to prevent future outbreaks.

## Data Availability

All data produced in the present study are available upon reasonable request to the corresponding author.

## Author Contributions

All the authors contributed to the intellectual content of this manuscript and approved the final draft. Conceptualisation, Yuki Takamatsu ; Methodology, Ryosaku Oshiro, Tadaki Suzuki, Akira Yoshikawa and Yuki Takamatsu ; Data collection, Ryosaku Oshiro, Motoki Ihara, Catarina Harumi Oda Ibrahim, Xayavong Dalouny, Vu Pham Hong Nhung, Yumika Takaki, and Hikaru Ogushi ; Investigation, Daichi Setoguchi, Shinichi Katsuoka, Tatsuki Murakami, Chikara Matsumoto, Akira Yoshikawa, and Yuki Takamatsu; Data analysis, Ryosaku Oshiro, Sho Miyamoto, Yusuke Sakai, Tadaki Suzuki, and Yuki Takamatsu; Writing – Original Draft Preparation, Ryosaku Oshiro, and Yuki Takamatsu; Writing – Review & Editing, Ryosaku Oshiro, Motoki Ihara, Catarina Harumi Oda Ibrahim, Kosuke Matsui, Xayavong Dalouny, Vu Pham Hong Nhung, Tomomi Kurashige, Qiang Xu, Shangfan Hu, Mya Myat Ngwe Tun, Yumika Takaki, Hikaru Ogushi, Daichi Setoguchi, Shinichi Katsuoka, Tatsuki Murakami, Chikara Matsumoto, Mitsuru Hattori, Naoko Hattori, Sho Miyamoto, Yusuke Sakai, Tadaki Suzuki, Hirotomo Yamanashi, Hiroyuki Murota, Takahiro Maeda, Kouichi Morita, Akira Yoshikawa, Yuki Takamatsu; Supervision, Yuki Takamatsu.

## Acknowledgments

The authors acknowledge Kazumi Jodai and Megumi Tsubota for their technical support, all members of the Department of Virology, Institute of Tropical Medicine, Nagasaki University, for their cooperation, and all medical staff at Goto Chuo Hospital for their care. This study was supported by the Japan Agency of Medical Research and Development (AMED) under grant numbers JP24fk0108656, JP24fk0108695, JP24wm0125006, JP24wm0125011, JP23fm0208101, JP23fk0108656, JP23wm0125006; the Japan Society for Promotion of Sciences under grant numbers, 22KK0115, 24K02288, the Asahi Glass Foundation, and Joint/Research Center on Tropical Disease, Institute of Tropical Medicine, Nagasaki University (2022-Ippan-12, 2023-Ippan-16); SCARDA project with JP223fa627004. The identified viruses will be provided by Nagasaki University through the National BioResource Project (Human pathogen viruses) of MEXT, Japan

## References

1. Karanam SK, Nagvishnu K, Uppala PK, et al. Crimean-Congo hemorrhagic fever: Pathogenesis, transmission and public health challenges. World J Virol 2025;14(1):100003. doi: 10.5501/wjv.v14.i1.100003

2. Chiffi G, Grandgirard D, Leib SL, et al. Tick-borne encephalitis: A comprehensive review of the epidemiology, virology, and clinical picture. Rev Med Virol 2023;33(5):e2470. doi: 10.1002/rmv.2470 [published Online : 20230701]

3. Seo JW, Kim D, Yun N, Kim DM. Clinical Update of Severe Fever with Thrombocytopenia Syndrome. Viruses 2021;13(7) doi: 10.3390/v13071213 [published Online : 20210623]

4. Park D, Kim KW, Kim YI, et al. Deciphering the evolutionary landscape of severe fever with thrombocytopenia syndrome virus across East Asia. Virus Evol 2024;10(1):veae054. doi: 10.1093/ve/veae054 [published Online : 20240720]

5. Mekata H, Umeki K, Yamada K, et al. Nosocomial Severe Fever with Thrombocytopenia Syndrome in Companion Animals, Japan, 2022. Emerg Infect Dis 2023;29(3):614–17. doi: 10.3201/eid2903.220720

6. Fu P, Meng Z, Peng Y, et al. Identification of severe fever with thrombocytopenia syndrome virus isolates in the northwest of Hubei Province, China. Acta Trop 2024;260:107397. doi: 10.1016/j.actatropica.2024.107397 [published Online : 20240914]

7. Jalal S, Hwang SY, Kim CM, et al. Comparison of RT-PCR, RT-nested PCRs, and real-time PCR for diagnosis of severe fever with thrombocytopenia syndrome: a prospective study. Sci Rep 2021;11(1):16764. doi: 10.1038/s41598-021-96066-4 [published Online : 20210818]

8. Yu XJ, Liang MF, Zhang SY, et al. Fever with thrombocytopenia associated with a novel bunyavirus in China. N Engl J Med 2011;364(16):1523–32. doi: 10.1056/NEJMoa1010095 [published Online : 20110316]

9. Takahashi T, Maeda K, Suzuki T, et al. The first identification and retrospective study of Severe Fever with Thrombocytopenia Syndrome in Japan. J Infect Dis 2014;209(6):816–27. doi: 10.1093/infdis/jit603 [published Online : 20131114]

10. Yoo JR, Heo ST, Park D, et al. Family Cluster Analysis of Severe Fever with Thrombocytopenia Syndrome Virus Infection in Korea. Am J Trop Med Hyg 2016;95(6):1351–57. doi: 10.4269/ajtmh.16-0527 [published Online : 20161017]

11. Yamanaka A, Kirino Y, Fujimoto S, et al. Direct Transmission of Severe Fever with Thrombocytopenia Syndrome Virus from Domestic Cat to Veterinary Personnel. Emerg Infect Dis 2020;26(12):2994–98. doi: 10.3201/eid2612.191513

12. Huang D, Jiang Y, Liu X, et al. A Cluster of Symptomatic and Asymptomatic Infections of Severe Fever with Thrombocytopenia Syndrome Caused by Person-to-Person Transmission. Am J Trop Med Hyg 2017;97(2):396–402. doi: 10.4269/ajtmh.17-0059 [published Online : 20170719]

13. Liu T, Zhang N, Li H, et al. Analysis of severe fever with thrombocytopenia syndrome cluster in east China. Virol J 2023;20(1):199. doi: 10.1186/s12985-023-02155-3 [published Online : 20230901]

14. Ishijima K, Taira M, Matsuu A, et al. 国 内 の ネ コ ・ イ ヌ に お け る 重症 熱 性 ⾎ ⼩ 板 減 少 症 候 群 の 発 ⽣ 状 況. Infectious Agents Surveillance Report 2023;44:31–32.

15. Ando T, Nabeshima T, Inoue S, et al. Severe Fever with Thrombocytopenia Syndrome in Cats and Its Prevalence among Veterinarian Staff Members in Nagasaki, Japan. Viruses 2021;13(6) doi: 10.3390/v13061142 [published Online : 20210614]

16. Osako H, Xu Q, Nabeshima T, et al. Clinical Factors Associated with SFTS Diagnosis and Severity in Cats. Viruses 2024;16(6) doi: 10.3390/v16060874 [published Online : 20240529]

17. Kobayashi Y, Kato H, Yamagishi T, et al. Severe Fever with Thrombocytopenia Syndrome, Japan, 2013-2017. Emerg Infect Dis 2020;26(4):692–99. doi: 10.3201/eid2604.191011

18. Matsuu A, Momoi Y, Nishiguchi A, et al. Natural severe fever with thrombocytopenia syndrome virus infection in domestic cats in Japan. Vet Microbiol 2019;236:108346. doi: 10.1016/j.vetmic.2019.06.019 [published Online : 20190720]

19. Sekizuka T, Itokawa K, Kageyama T, et al. Haplotype networks of SARS-CoV-2 infections in the Diamond Princess cruise ship outbreak. Proc Natl Acad Sci U S A 2020;117(33):20198–201. doi: 10.1073/pnas.2006824117 [published Online : 20200728]

20. Ishikawa F, Udaka Y, Oyamada H, et al. Genetic epidemiology using whole genome sequencing and haplotype networks revealed the linkage of SARS-CoV-2 infection in nosocomial outbreak. Infect Prev Pract 2021;3(4):100190. doi: 10.1016/j.infpip.2021.100190 [published Online : 20211124]

21. Shimada S, Aoki K, Nabeshima T, et al. Tofla virus: A newly identified Nairovirus of the Crimean-Congo hemorrhagic fever group isolated from ticks in Japan. Sci Rep 2016;6:20213. doi: 10.1038/srep20213 [published Online : 20160211]

22. Sun J, Qian L, Li D, et al. Concurrent severe fever with thrombocytopenia syndrome virus outbreaks on multiple fox farms, China, 2023. Emerg Microbes Infect 2025;14(1):2447610. doi: 10.1080/22221751.2024.2447610 [published Online : 20250112]

